# Sex disparities in COVID-19 mortality vary considerably across time: The case of New York State

**DOI:** 10.1101/2022.01.11.22269104

**Authors:** Ann Caroline Danielsen, Marion Boulicault, Annika Gompers, Tamara Rushovich, Katharine MN Lee, Sarah S. Richardson

**Affiliations:** Harvard GenderSci Lab, Cambridge, MA, USA; Department of Linguistics and Philosophy, Massachusetts Institute of Technology, Cambridge, MA, USA; Department of Obstetrics and Gynecology, Beth Israel Deaconess Medical Center, Boston, MA, USA; Population Health Sciences Department, Harvard Graduate School of Arts and Sciences, Cambridge, MA, USA; Department of Social and Behavioral Sciences, Harvard T.H. Chan School of Public Health, Boston, MA, USA; Division of Public Health Sciences, Department of Surgery, Washington University in St Louis School of Medicine, St Louis, MO; Studies of Women, Gender, and Sexuality, Harvard University, Cambridge, MA, USA; Department of the History of Science, Harvard University, Cambridge, MA, USA

**Author notes:** **Corresponding author** Ann Caroline Danielsen.

**Keywords:** COVID-19, sex disparities, temporal analysis

## Abstract

In order to characterize how sex disparities in COVID-19 mortality evolved over time in New York State (NY), we analyzed sex-disaggregated data from the US Gender/Sex COVID-19 Data Tracker from March 14, 2020 to August 28, 2021. We defined six different time periods and calculated mortality rates by sex and mortality rate ratios, both cumulatively and for each time period separately. As of August 28, 2021, 19 227 (44.2%) women and 24 295 (55.8%) men died from COVID-19 in NY. 72.7% of the cumulative difference in the number of COVID-19 deaths between women and men was accrued between March 14 and May 4, 2020. During this period, the COVID-19 mortality rate ratio for men compared to women was 1.56 (95% CI: 1.52-1.61). In the five subsequent time periods, the corresponding ratio ranged between 1.08 (0.98-1.18) and 1.24 (1.15-1.34). While the cumulative mortality rate ratio of men compared to women was 1.34 (1.31-1.37), the ratio equals 1.19 (1.16-1.22) if deaths during the initial COVID-19 surge are excluded from the analysis. This article shows that in NY the magnitude of sex disparities in COVID-19 mortality was not stable across time. While the initial surge in COVID-19 mortality was characterized by stark sex disparities, these were greatly attenuated after the introduction of public health controls.

## Introduction

Men in aggregate have died from COVID-19 at higher rates than women, with initial assessments claiming that men were dying at twice the rate of women.^1^ This sex disparity is frequently assumed to remain stable over time and across contexts, supporting a primary role for biological sex-related variables in men’s greater susceptibility to severe COVID-19.^2^ No studies have examined the variation of sex disparities in COVID-19 mortality over time. Attending to how mortality patterns vary across time, however, uncovers dynamics that are otherwise obscured and reveals how sociocontextual factors may contribute to sex disparities in infectious disease outbreaks.^3^

Here we examine the case of New York State (NY), a global index site in the COVID-19 pandemic.^4^ New York (NY) is in the top quintile for cumulative sex disparity in COVID-19 mortality in the United States.^5^ Further, despite accounting for 6% of the total US population, as of September 2021 NY has contributed 8% of the country’s COVID-19 deaths and 9% of total excess male deaths.^6,7^ Below, we describe large changes in mortality by sex across different phases of the pandemic in NY.

## Methods

We obtained weekly sex-disaggregated COVID-19 mortality data for NY from the US Gender/Sex COVID-19 Data Tracker from March 14, 2020 through August 28, 2021.^5^ Fatality data reported by the Tracker include all individuals categorized as women or men who died from COVID-19 in NY, as reported by the New York State Department of Health.^8^ Data used in this analysis were publicly available and de-identified and are exempt from IRB oversight.

Mortality rates by sex were calculated by dividing the total number of deaths in each sex stratum by the corresponding total population residing in NY. The 2015-2019 5-Year American Community Survey population estimates for NY served as sex-disaggregated population denominators.^9^ To capture changes over time, mortality rates by sex were calculated cumulatively over the total period of observation and separately for six different time periods across the waves of the pandemic and phases of the vaccination campaign in NY. Rates are reported per 100 000 people. To compare mortality among women and men, rate ratios were computed for each time period, taking women as the reference population. Deaths with sex categorized as “unknown” or “other” (making up 0.03% of the data) were excluded from the analysis.

The six time periods were defined as follows:

1. March 14, 2020 to May 4, 2020

2. May 5, 2020 to June 1, 2020

3. June 2, 2020 to October 26, 2020

4. October 27, 2020 to December 14, 2020

5. December 15, 2020 to April 5, 2021

6. April 6, 2021 to August 28, 2021

## Results

A total of 43 522 individuals categorized as women or men are recorded as having lost their lives to COVID-19 between March 14, 2020 and August 28, 2021 in NY and were included in the study. Of these, 19 227 (44.2%) are women and 24 295 (55.8%) are men (Table 1). The cumulative mortality rate was 190.91 per 100 000 (95% CI: 188.21-193.61) for women and 255.71 (252.49, 258.92) for men, corresponding to a mortality rate ratio of 1.34 (1.31-1.37).

**Table 1:** COVID-19 mortality rates and rate ratios in New York State in different time periods, disaggregated by sex. *Rate per 100 000 people. ** Rate ratios are calculated with women as the reference population.

|  | Count (%) |  | Rate* (95% CI) |  | Rate Ratio** |
| --- | --- | --- | --- | --- | --- |
|  | Women | Men | Women | Men |  |
| Mortality by Time Period |  |  |  |  |  |
| 14 Mar 2020<br>- 4 May 2020 | 7 747 (40.4) | 11 433 (59.6) | 76.92 (75.21,<br>78.63) | 120.33<br>(118.13,<br>122.54) | 1.56 (1.52,<br>1.61) |
| 5 May 2020 -<br>1 Jun 2020 | 2 326 (49.3) | 2 390 (50.7) | 23.10 (22.16,<br>24.03) | 25.16 (24.15,<br>26.16) | 1.09 (1.03,<br>1.15) |
| 2 Jun 2020 -<br>26 Oct 2020 | 905 (49.6) | 920 (50.4) | 8.99 (8.40,<br>9.57) | 9.68 (9.06,<br>10.31) | 1.08 (0.98,<br>1.18) |
| 27 Oct 2020 -<br>14 Dec 2020 | 996 (48.5) | 1,059 (51.5) | 9.89 (9.26,<br>10.50) | 11.15 (10.47,<br>11.82) | 1.13 (1.03,<br>1.23) |
| 15 Dec 2020 -<br>5 Apr 2021 | 5 974 (46.1) | 6 996 (53.9) | 59.32 (57.81,<br>60.82) | 73.63 (71.91,<br>75.36) | 1.24 (1.20,<br>1.28) |
| 6 Apr 2021 -<br>28 Aug 2021 | 1 279 (46.1) | 1 497 (53.1) | 12.70 (12.00,<br>13.40) | 15.76 (14.96,<br>16.55) | 1.24 (1.15,<br>1.34) |
| Cumulative Mortality |  |  |  |  |  |
| 14 Mar 2020<br>- 28 Aug 2021 | 19 227 (44.2) | 24 295 (55.8) | 190.91<br>(188.21,<br>193.61) | 255.71<br>(252.49,<br>258.92) | 1.34 (1.31,<br>1.37) |

| Cumulative Mortality after 4 May, 2020 |  |  |  |  |  |
| --- | --- | --- | --- | --- | --- |
| 5 May 2020 -<br>28 Aug 2021 | 11 480 (47.2) | 12 862 (52.8) | 113.99<br>(111.90,<br>116.07) | 135.38<br>(133.04,<br>137.71) | 1.19 (1.16,<br>1.22) |

Notably, 3,686 (72.7%) of the 5 068 excess deaths among men took place between March 14 and May 4, 2020 (Figure 1). During this period, the mortality rate among women was 76.92 (75.21-78.63), while the mortality rate among men was 120.33 (118.13-122.54). The corresponding mortality rate ratio for men compared to women in this period was 1.56 (1.52-1.61).

**Figure 1:**
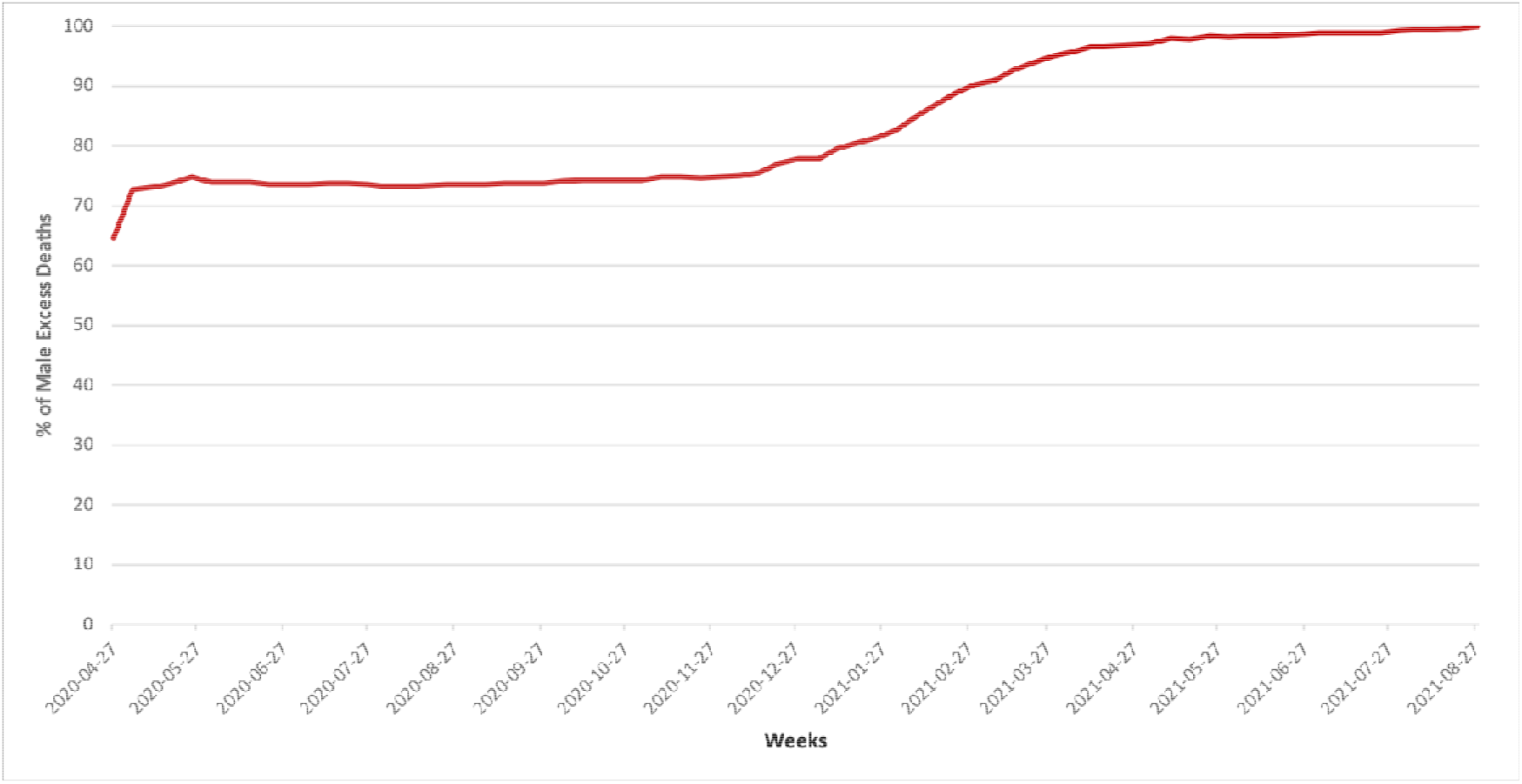
Proportion of the total difference between the number of male and female deaths that was accrued week by week between March 14, 2020 and August 28, 2021.

Our analysis reveals that sex disparities changed drastically during subsequent phases of the pandemic. Between May 5 and June 1, 2020, the mortality rate among women and men was respectively 23.10 (22.16-24.03) and 25.16 (24.15-26.16), corresponding to a rate ratio of 1.09 (1.03-1.15). The sex disparity showed a similar pattern between June 2 and October 26, 2020, with the mortality rate ratio of men compared to women equalling 1.08 (0.98-1.18), and remained stable through December 14, 2020 (mortality rate ratio: 1.13; 1.03-1.23). During the subsequent period (December 15 2020 to April 5, 2021), we observe an increase in mortality rates for both women (59.32; 95% CI: 57.81-60.82) and men (73.63; 71.91-75.36), resulting in a rate ratio of 1.24 (1.20-1.28). Despite mortality rates decreasing for both women and men between April 6 and August 28, 2021 compared to the previous period, the rate ratio remained 1.24 (1.15-1.34). If fatalities between March 14 and May 4, 2020 are excluded from the analysis, the mortality rate was 113.99 (111.90-116.07) for women and 135.38 (133.04-137.71) for men, corresponding to a mortality rate ratio of 1.19 (1.16, 1.22).

## Discussion

Using unique sex-disaggregated longitudinal data from the US Gender/Sex COVID-19 Data Tracker, our analysis shows significant variation in the magnitude of sex disparities in COVID-19 mortality in NY over time. The sex disparity was greatest early in the pandemic between March 14 and May 4, 2020, when the mortality rate among men was 1.56 (95% CI: 1.52-1.61) times the mortality rate among women. Despite the fact that only 44.1% of total deaths in the state occurred in this period, the period accounts for 72.7% of the total difference in the number of deaths between women and men, creating a gap in mortality rates that continues to affect the cumulative sex disparity to the present day.^5^ While the cumulative mortality rate for men compared to women across all periods is 1.34 (1.31-1.37), the rate decreases to 1.19 (1.16-1.22) if the initial period is excluded from the analysis (May 5, 2020 to August 28, 2021). This ratio should be contextualized, for example, against the 1.58 mortality rate ratio of Nevada, a state that implemented few public health controls over the course of the pandemic.^5^

Many gender-related social and demographic factors could affect variation in the sex disparity over time.^10–12^ Of note, New York City (NYC) accounts for 80.7% of male excess deaths in NY over the entirety of the pandemic, while only accounting for 53.1% of all COVID-19 deaths in the state.^7^ As is well known, NY and in particular NYC were hit by COVID-19 especially early and severely compared to the rest of the US. During this time, NY instituted several provisions to curb the spread of the pandemic. For example, non-essential businesses were closed on March 22, individuals were banned from gathering on March 24, and face masks were required in public places beginning April 17.^13^ The precipitous decline in male mortality observed after May 4, 2020 follows the progressive implementation of pandemic-control provisions, thereby supporting existing research suggesting that gendered behavioral, occupational, and structural factors play a central role in determining disparities in COVID-19 mortality.^10,11,14^

The relative increase in male mortality during the fifth and sixth periods coincided with the second major COVID-19 surge in NY, as well as the large-scale roll-out of COVID-19 vaccines, with the first vaccine being administered in NY on December 15, 2020 and all individuals older than 16 years becoming vaccine-eligible on April 6, 2021. To date, women account for the majority of vaccine recipients in NY.^15^ Therefore, gender/sex patterns of vaccination linked to age, demographics, occupation, health behaviors, and other social variables are likely affecting COVID-19 disparities in the context of the gradual relaxation of pandemic provisions during the final time periods in our study.

Available sex-disaggregated COVID-19 mortality data in NY are not reported in conjunction with any other demographic variables. Data on sex as it interacts with other factors such as age, race/ethnicity, socioeconomic status, occupation, and comorbidity are crucial to better understanding sex disparities in COVID-19.^10,12^ For example, Rushovich et al. showed that aggregate sex comparisons obscured very high COVID-19 fatality rates for black women compared to both white women and white men, and relatively low fatality rates for white men, compared to black men.^10^

Overall, these findings demonstrate that in New York State, an early and severely impacted global index site in the COVID-19 pandemic, sex disparities in COVID-19 mortality have not remained stable across time and were greatly attenuated after the initial, most acute and deadly phase, prior to the introduction of public health controls. Our findings suggest a critical role for gendered behavioral, occupational, and structural factors in driving sex disparities in infectious disease outbreaks and underscore the importance of investigating temporal changes in the magnitude of sex disparities for understanding and addressing their causes.^16^

### Implications for Policy and Practice

- The substantial attenuation in male COVID-19 mortality after the initial spring 2020 surge in New York State supports an important role for public health controls in protecting men in infectious disease epidemics.
- This article argues that cumulative estimates of sex disparity in COVID-19 mortality are misleading and hinder opportunities to evaluate the relationship between sex disparities and public health controls.
- Investigating how sex disparities in COVID-19 have changed over time is critical for illuminating and addressing their root causes.
- This article demonstrates the need for further research investigating the relationship between changes in sex disparity magnitude over time and gendered behavioral, occupational, and structural factors.

## Data Availability

Original data used for these analyses are available online as part of the US Gender/Sex COVID-19 Data Tracker

https://www.genderscilab.org/gender-and-sex-in-covid19

## Acknowledgments

Thank you to Capri D’Souza, Mimi Tarrant, Kai Jillson, May Moorefield and Kashfia Rahman for their work collecting and validating data for the Gender/Sex COVID-19 Data Tracker, and to members of the Harvard GenderSci Lab for their feedback on earlier drafts of the manuscript.

## References

1. Baker P, White A, Morgan R. Men’s health: COVID-19 pandemic highlights need for overdue policy action. The Lancet. 2020;395(10241):1886–1888. doi:10.1016/S0140-6736(20)31303-9

2. Scully EP, Haverfield J, Ursin RL, Tannenbaum C, Klein SL. Considering How Biological Sex Impacts Immune Responses and COVID-19 Outcomes. Nature Reviews Immunology. 2020;20(7):442–447. doi:10.1038/s41577-020-0348-8

3. Krieger N, Chen JT, Waterman PD. Excess mortality in men and women in Massachusetts during the COVID-19 pandemic. The Lancet. 2020;395(10240):1829. doi:10.1016/S0140-6736(20)31234-4

4. Lajous M, Huerta-Gutiérrez R, Kennedy J, Olson DR, Weinberger DM. Excess Deaths in Mexico City and New York City During the COVID-19 Pandemic, March to August 2020. Am J Public Health. 2021;111(10):1847–1850. doi:10.2105/AJPH.2021.306430

5. US Gender/Sex COVID-19 Data Tracker. GenderSci Lab. Published 2020. Accessed September 30, 2021. https://www.genderscilab.org/gender-and-sex-in-covid19

6. US Census Bureau. US Census Bureau QuickFacts. Accessed September 30, 2021. https://www.census.gov/quickfacts/fact/table/US/PST045219

7. National Center for Health Statistics. Provisional COVID-19 Death Counts by Sex, Age, and State. Centers for Disease Control and Prevention. Published 2021. Accessed September 29, 2021. https://data.cdc.gov/NCHS/Provisional-COVID-19-Death-Counts-by-Sex-Age-and-S/9bhg-hcku

8. New York State Department of Health. NYS COVID-19 Tracker. NYS COVID-19 Tracker. Published 2020. Accessed September 30, 2021. https://covid19tracker.health.ny.gov/views/NYS-COVID19-Tracker/NYSDOHCOVID-19Tracker-Fatalities?%3Aembed=yes%3Atoolbar=no%3Atabs=n

9. US Census Bureau. American Community Survey, 2015-2019 American Community Survey 5-Year Estimates, Table B01001; generated by Tamara Rushovich; using tidycensus in R. The United States Census Bureau. Published April 12, 2021. Accessed April 21, 2021. https://www.census.gov/data/developers/data-sets/acs-5year.html

10. Rushovich T, Boulicault M, Chen JT, et al. Sex Disparities in COVID-19 Mortality Vary Across US Racial Groups. J GEN INTERN MED. 2021;36(6):1696–1701. doi:10.1007/s11606-021-06699-4

11. Hawkins D. Differential occupational risk for COVID-19 and other infection exposure according to race and ethnicity. American Journal of Industrial Medicine. 2020;63(9):817–820. doi:https://doi.org/10.1002/ajim.23145

12. Chen JT, Krieger N. Revealing the Unequal Burden of COVID-19 by Income, Race/Ethnicity, and Household Crowding: US County Versus Zip Code Analyses. Journal of Public Health Management and Practice. 2021;27:S43. doi:10.1097/PHH.0000000000001263

13. Francescani C. Timeline: The first 100 days of New York Gov. Andrew Cuomo’s COVID-19 response. ABC News. https://abcnews.go.com/US/News/timeline-100-days-york-gov-andrew-cuomos-covid/story?id=71292880. Published June 17, 2020.

14. Griffith DM, Sharma G, Holliday CS, et al. Men and COVID-19: A Biopsychosocial Approach to Understanding Sex Differences in Mortality and Recommendations for Practice and Policy Interventions. Prev Chronic Dis. 2020;17. doi:10.5888/pcd17.200247

15. New York State. Vaccine Demographic Data. COVID-19 Vaccine. Accessed September 30, 2021. https://covid19vaccine.health.ny.gov/vaccine-demographic-data

16. Riley AR. Advancing the study of health inequality: Fundamental causes as systems of exposure. SSM Popul Health. 2020;10. doi:10.1016/j.ssmph.2020.100555

